# Microbiome and Genetic Predictors of Weight Loss 12 Months Post Sleeve Gastrectomy: Insights from a Pilot Retrospective Cohort Study

**DOI:** 10.1101/2025.01.14.25319888

**Authors:** Inti Pedroso, Shreyas V. Kumbhare, Shaneeta Johnson, Karthik M. Muthukumar, Santosh K. Saravanan, Carmel Irudayanathan, Garima Sharma, Lawrence Tabone, Ranjan Sinha, Daniel E. Almonacid, Nova Szoka

## Abstract

**Background:** Gut microbiome and genetic biomarkers are increasingly guiding obesity treatment. Bariatric surgery leads to shifts in gut microbial composition and function, while genome-wide association studies reveal genetic underpinnings of polygenic obesity, informing risk, therapeutic outcomes, and nutrigenomics-based interventions.

**Objectives:** This pilot study aimed to identify gut microbiome and genetic biomarkers associated with weight loss 12 months after sleeve gastrectomy (SG).

**Setting:** Single academic institution university clinic

**Methods:** Sixty-seven patients 12 months post-SG were enrolled: 34 had successful excess weight loss (EWL≥50%), while 33 had unsuccessful EWL (EWL<50%). Microbiome and genetic profiles were collected and analyzed using ANOVA and regression methods.

**Results:** The genus Akkermansia was significantly associated with EWL (p=9.9×10^−6). Several microbial pathways, including propionate synthesis and menaquinone (vitamin K2) production, showed nominally significant (p<0.05) associations with greater weight loss. No differences emerged in the Firmicutes/Bacteroidetes ratio. Genetic analyses revealed significant correlations between EWL and polygenic scores for dietary needs and metabolic responses, including distinct vitamin D and K requirements, as well as higher LDL cholesterol levels and predisposition for major depression.

**Conclusion:** These findings suggest that both the gut microbiome and genetics may modulate weight loss following bariatric surgery. Integrating microbiome and genetic profiling into bariatric care pathways could enhance personalized obesity treatment. While this pilot, exploratory, and proof-of-concept study has limitations, it supports prior work linking gut microbial pathways to weight loss and suggests new associations. Follow-up studies are warranted to validate these results and further inform precision obesity interventions.

**Highlights:**

- Akkermansia strongly correlates with weight loss 12 months post-sleeve gastrectomy.
- Microbiome propionate and vitamin K2 pathways linked to higher excess weight loss.
- Genetic markers link vitamin needs, LDL and depression risk with lower weight loss.
- Microbiome and genetics integration may improve bariatric surgery outcomes.

## Introduction

Obesity is a complex metabolic disease and increasing global epidemic, as well as a risk factor for diabetes, cardiovascular disease, and reduced life expectancy, that currently affects 42% of the adult population in the United States [1]. Bariatric surgery is the most effective intervention for morbid obesity and outcomes are intricately intertwined with physiologic, behavioral, and psychological factors [2].

Research indicates that individuals’ gut microbiome and genetic makeup are intrinsically linked to their metabolism [3,4]. The gut microbiota is implicated in obesity through its ability to influence energy metabolism, appetite regulation, and nutrient absorption [4].; Imbalances in microbial composition can lead to increased energy harvest from foods and alterations in metabolic processes that contribute to weight gain; thus, individuals with obesity display an altered gut microbiome [4]. Bariatric surgery leads to significant changes in the gut microbiome, increasing microbial gene richness and shifting toward a “lesser obese” microbial structure [5]. Furthermore, changes in the gut microbiome persist up to a decade after bariatric surgery compared to individuals with severe obesity who do not undergo surgery [6]. In the past, the genetics of obesity led to disease classification as monogenic versus polygenic (or common) obesity, however recent genome-wide association studies (GWAS) suggest that both forms of obesity have shared genetic foundations and implicate the central nervous system leptin-melanocortin pathway as a major driver of body weight regulation [3,4].

The growing prevalence of obesity necessitates a shift toward multimodal treatment strategies that integrate both traditional and innovative interventions to address its multifaceted nature, shaped by behavioral, metabolic, and genetic factors. Precision obesity care, leveraging microbiome profiling and genetic testing, offers promising avenues for creating targeted therapeutic and nutritional strategies. This comprehensive, personalized, precision-based approach mirrors advances in other fields like oncology, where integrated multimodal care has improved outcomes, and provides a framework for optimizing obesity management and enhancing patient outcomes [7].

In this pilot proof of concept study, we utilized microbiome and genetic profiling to investigate genetics and microbiome correlates of weight loss 12 months post-sleeve gastrectomy (SG) to identify biomarkers that can inform personalized treatment plans.

## Methods

### Study Design and Recruitment

This study was performed under IRB Approval (WVU Protocol #2006039647). Adult patients who underwent sleeve gastrectomy (SG) at West Virginia University and were 12 months post-surgery were recruited from the bariatric clinic from 12/1/2020 through 11/30/2022. Patients age 18 and older were deemed eligible, and recruitment occurred at their routine 12 month post operative follow up visit. Exclusion criteria were nonbariatric patients, patients who had their initial bariatric operation at an institution other than WVU, patients who underwent a gastric bypass surgery or who had who had a gastric band with subsequent revisional surgery in the WVU Bariatric program, patients who lack resources to complete remote coaching sessions, and vulnerable patient populations, namely pregnant and lactating women, prisoners, cognitively impaired and critically ill subjects. All patients with inclusion criteria were offered to participate and 67 accepted to enroll. Successful weight loss after SG is clinically defined as achieving at least 50% excess weight loss (EWL), calculated as %EWL = 100*[(start_BMI - 25) - (end_weight - 25)]/(start_BMI - 25), with BMI being Body Mass Index [8]. We sought to identify multi-omic correlates of EWL 12 months post-sleeve gastrectomy. After recruitment at WVU the study subjects were referred to and enrolled in the Digbi Health personalized care program for weight loss. Upon enrollment, patients provided self-reported demographic and health information through online surveys and genetic and gut microbiome samples using at home collection kits. Patients were provided with at-home sample collection kits within the first week after enrolling with Digbi Health together with detailed printed instructions and support from a trained coach for at-home sample collection. All individuals were reminded to return their samples as soon as possible and in biweekly intervals afterwards. Individuals that failed to return the samples after 6 weeks from receiving them were deemed as missing data. Research associated with this weight loss program has been previously reviewed and approved by the Institutional Review Board of E&I Review Services (protocol code #18053 on 05/22/2018). All subjects considered for the present study provided their research informed consent electronically as part of their written informed consent form.

### Sample collection and processing

Subjects self-collected gut microbiome (fecal) samples using fecal swabs (Mawi Technologies iSWAB Microbiome collection kit, Model no. ISWAB-MBF-1200) and saliva samples using buccal swabs, following standardized protocols. Fecal DNA extraction was performed using Qiagen MagAttract Power Microbiome DNA Kit on an automated liquid handling DNA extraction instrument, followed by bacterial 16S rRNA gene V3-V4 region amplification and sequencing on the Illumina MiSeq platform using 2 × 300 bp paired-end sequencing performed at Akesogen Laboratories in Atlanta, GA. Sequence reads were demultiplexed, denoised and ASVs generated using DADA2 in QIIME2 version 2021.4 [9]. DNA genotyping was conducted using Affymetrix’s Direct-to-Consumer Array version 2.0 on the GeneTitan platform at Akesogen Laboratories in Atlanta, GA, with genotype calling and data analysis performed according to established best practices detailed in prior research [10].

### Microbiome and genetic data analysis

Initial quality control steps included the removal of primers and low-quality bases, removing ASVs classified as non-bacterial sequences (such as Euryarchaeota, Chloroplast, Mitochondria), or unassigned taxa at the phylum level. Taxa were agglomerated at genus levels, and those with low abundance (taxa with <10 reads in at least 10% of samples) were excluded, resulting in a reduction of the sparsity of the abundance matrix to 41.57% and removal of singletons. The abundance matrix was rarefied at even depth (n=73,000 reads per sample (minimum reads across the samples) with 500 iterations) using QIIME2, resulting in 177 taxa across 39 samples. The abundance of functional microbial pathways related to gut and neuroactive metabolites was calculated as described elsewhere [11]. All raw abundances were centered-log ratio (CLR) transformed unless otherwise specified.

For DNA profiling, probe-level DNA genotype call files were obtained from the genotyping facility and formatted in VCF format with QC steps including removal of discordant genotypes and left normalization. Beagle version 5.3 [12,13] was used for phasing and imputation using the 1KG project as reference panel [14] and genetic variants with imputation r^2^≥0.8 were selected for downstream analyses. We computed polygenic scores for a set of selected phenotypes relevant to nutritional guidance, obesity, diabetes, and comorbid conditions. To this end variants effect sizes were obtained from published articles, supplementary materials and GWAS [15] or PGS Catalog [16] databases. Table S1 provides a list of references to the original underlying each PGS calculated. PGS were calculated using PLINK2 “plink2 --score” command [17]. All genetic scores were coded to be interpreted such that a larger genetic score is associated with increasing inherited genetic predisposition to the condition.

### Statistical and data analysis

We used PERMANOVA to perform community-level (microbiome) multivariate association with variables, based on the abundance matrix using the Bray-Curtis dissimilarity, and using the “vegan” package (“adonis2” function) for R. To further test for homogeneity of multivariate dispersions (comparing inter-individual variations) between groups, we used the betadisper test. To identify taxa and functional pathways associated with EWL we utilized linear models implemented on the GAMLSS software package for R [18]. We used the following regression formulas to test the association with EWL: “abundance∼1 + Gender + Age + Initial_BMI + perc_EWL” and with the following options ‘gamlss(…., control=gamlss.control(c.crit = 0.001, n.cyc = 2000), family = BEZI())’, where “abundance” corresponds to the rarified counts of the taxa or functions divided by the total counts of the sample, “Initial_BMI” is the BMI at the time of enrollment at Digbi and included in the regression due to its known relationship to the gut microbiome, and “BEZI” is the beta distribution family (Beta Zero Inflated). We performed corrections for multiple hypothesis tests using the local FDR (lfdr) methodology implemented on the “ashr” software package for R [19], using the beta and standard error obtained in the regression analysis and with the option “mixcompdist=‘normal’”. We set a local lfdr of 0.15 as the cutoff to declare statistical significance considering that this is a proof-of-concept study aiming to identify multi-omic associations warranting follow-up in studies with better statistical power. For microbiome analyses, we excluded from the regression analyses, the genera found in less than 20 individuals (with relative abundance > 0) to avoid false positives due to smaller sample sizes. The differences between groups for Firmicutes/Bacteroidetes ratio were tested using ANOVA. To test for the association between PGS and EWL we used the same strategy described for microbiome except that we excluded “Initial_BMI” from the regression.

## Results

### Cohort characteristics

Sixty-seven adult patients who were 12 months post sleeve gastrectomy were recruited at our academic bariatric clinic from 12/1/2020 through 11/30/2022. Of these, 52 participants enrolled in the Digbi Health dietary intervention for weight loss. Forty-two participants remained engaged for more than 4 months and 39 provided comprehensive weight, health, and lifestyle data, along with genetic and gut microbiome samples (Table S2). 34 participants had a EWL ≥ 50%, while 33 participants had a EWL<50% (Figure S1). There were no significant differences between groups in terms of age (EWL> ≥ 50% = 45.4 ± 9.82, EWL < 50% = 47.5 ± 10.37; p= 0.515) and gender (EWL ≥ 50%: Male= 2, Female = 18; EWL < 50%: Male= 4, Female = 16; p=0.658).

### Association of gut microbiome markers with excess weight loss

We first evaluated the overall association between the gut microbiome profiles and the EWL health outcome, and this association did not reach statistical significance (p = 0.06, Figure S2A). Guided by an association previously reported in the literature we tested the Firmicutes/Bacteroidetes for its association with EWL and found no association (p = 0.80, Figure S2B).

To test the association of individual gut microbiome markers, we analyzed 106 gut microbiome genera for their associations with EWL of which 6 had a nominally significant association (p < 0.05) and one, Akkermansia (p = 9.98×10^−6^ and lfdr = 5.38×10^−4^), reached statistical significance after multiple testing correction (lfdr < 0.15) (see Table 1 and Table S3). We also tested the association of 140 gut microbial pathways, identifying 8 with nominally significant associations (p < 0.05). These did not reach statistical significance (lfdr > 0.15) (see Table S4). These results indicate that individuals achieving EWL ≥ 50% 12 months post-SG, had a gut microbiome with a higher abundance of Akkermansia, and pathways associated with propionate synthesis (MGB054, p-value = 0.0095), menaquinone (vitamin K2) production (MGB041, p-value = 0.0276), and fucose degradation (MF0016, p-value = 0.0088), alongside other metabolic pathways such as arabinoxylan (MF0001, p-value = 0.0280) and arabinose degradation (MF0014, p-value = 0.0378), lactate consumption (MF0080, p-value = 0.0273), and sulfate reduction (MF0102, p-value = 0.0375) (Table S4). Table S5 and S6 provide abundance for genera and microbial pathways reaching nominally significant associations. Figures S3 to S8 and Figures S9 to S17 provide boxplots as a graphical representation of these association’s results.

### Association of genetic markers with excess weight loss

We analyzed 107 polygenic scores (PGS) for their associations with EWL outcomes 12 months post-SG, and identified six associations reaching nominal statistical significance (p < 0.05) but none reaching significance after multiple testing corrections (lfdr > 0.15) (see Table S7). These results indicate that those achieving EWL ≥ 50% exhibited a higher propensity to higher Vitamin K needs (p-value = 0.0011), lower Vitamin D needs (p-value = 0.0006), and lower predisposition to Major Depression (p-value = 0.02), Psoriasis (p-value = 0.027), high LDL Cholesterol levels (p-value = 0.043) and Protein Glycation (p-value = 0.046) levels. Table S8 provides PGS values for the scores reaching nominally significant associations. Figures S18 to S23 provide boxplots as a graphical representation of these associations’ results.

### Genetics and Microbiome interactions effects on excess weight loss

We noted that both the genetic and gut microbiome data provided marginally significant association with vitamin K metabolism and, therefore, we evaluated a model to see if there exists a synergistic interaction between genetics and microbiome components of the Vit K metabolism on the EWL. The model including genetic and microbiome variables corroborated a marginally significant association for each component separately (PGS Vit-K needs p-value=0.023 and Microbiome MGB04 p-value=0.04) but did not find statistical evidence of association for the interaction between the two (interaction p-value = 0.15).

## Discussion

Our study investigated the associations between genetic predispositions, gut microbial genera, and pathways, with EWL outcome in a cohort of 39 individuals, 12 months post-sleeve gastrectomy. We identified a statistically significant association between the gut microbiome genus *Akkermansia* and EWL. There was no association of the Firmicutes/Bacteroidetes ratio with EWL, which supports earlier findings by Duncan et al. [20]. While no significant differences were observed in the microbial diversity and composition, we found a nominally significant association between gut microbial pathways such as propionate synthesis and simple sugar metabolism and weight loss. Our findings identified associations between PGS traits such as vitamin D and K needs, predisposition to major depression, and high LDL cholesterol levels, underscoring the potential role of genetics in influencing weight loss success and obesity related comorbidity resolution.

The statistically significant association between the genus *Akkermansia* (p = 9.98×10^−6^ and lfdr = 5.38×10^−4^) and EWL is consistent with previous studies which showed that bariatric surgery significantly alters the gut microbiota, increasing microbial diversity, including the relative abundance of *Akkermansia* [21]. The impact of bariatric surgery on the gut microbiome and *A. muciniphila* has been shown to persist for up to a decade [6]. Importantly, the increased abundance of *A. muciniphila* has been linked to improvements in gut barrier function, reduced intestinal permeability, and metabolic regulation, which collectively support better weight loss outcomes. We and others have previously shown [22,23] that a microbiome-targeted dietary intervention can systematically support a higher abundance of *A. muciniphila* suggesting that this strategy may be a promising alternative to support better health outcomes in bariatric surgery patients.

The analysis of gut microbial pathways revealed associations with glucose metabolism, including propionate synthesis, fucose degradation, and menaquinone (vitamin K2) production. Although these did not remain significant after multiple testing corrections. Propionate, a short-chain fatty acid produced through the fermentation of dietary fibers, plays a critical role in metabolic health by stimulating satiety hormones such as peptide YY (PYY) and glucagon-like peptide-1 (GLP-1), which reduce appetite and promote feelings of fullness [24]. Additionally, propionate has been shown to improve insulin sensitivity and inhibit hepatic lipid synthesis, contributing to better glucose regulation and reduced fat accumulation [24]. Similarly, menaquinone (vitamin K2), synthesized by gut bacteria, is recognized for its role in calcium homeostasis and insulin sensitivity [25] by regulating vitamin K-dependent proteins like osteocalcin, which modulate energy metabolism and fat storage [26]. Its anti-inflammatory properties may also mitigate chronic inflammation, a common driver of obesity and insulin resistance. These findings suggest that microbial pathways such as propionate synthesis and menaquinone production could serve as promising biomarkers and mediate weight loss success, offering potential targets for personalized interventions.

The results of the PGS analysis showed associations between genetic traits and EWL outcomes 12 months post-sleeve gastrectomy (SG). EWL was associated with lower vitamin D needs and higher vitamin K requirements, highlighting the role of genetic factors in modulating nutrient metabolism and influencing post-surgical outcomes. These findings align with existing evidence suggesting that polygenic scores for vitamin D metabolism can affect weight loss outcomes, particularly when interacting with vitamin D levels and diet-related factors [27,28]. Prior studies have shown that genetic variations, such as in the vitamin D receptor (VDR), influence responses to weight loss interventions and that vitamin D sufficiency can mitigate genetic obesity risk [27]. These observations may motivate precision nutrition interventions that integrate genetic profiles to optimize dietary recommendations to enhance post-surgical weight loss.

Our results point towards an association between genetic predisposition to major depression and reduced EWL. Although not statistically significant after multiple testing corrections, it is aligned with previous findings indicating that both genetic predisposition to and depressive symptoms are associated with lower EWL after bariatric surgery potentially mediated by exacerbated peripheral inflammation and or psychological factors [29,30].

Interestingly, Vitamin K2 metabolism was associated with the genetic and gut microbiome data and in-spite of not finding evidence of a synergistic interaction between the two, our results suggest that both may be considered additively when personalizing an individual’s dietary intake to improve weight loss outcomes.

This study has several limitations that must be acknowledged. Its relatively small sample size and limited statistical power restrict the robustness of our findings. As a single-institution pilot study, the generalizability of the findings is restricted, and larger, multi-center studies will be required to validate and expand these results. Furthermore, the exploratory nature of these analyses highlights the need for cautious interpretation, as these findings serve primarily to generate hypotheses for future research. Despite these limitations, the study’s results replicated and expounded on previous findings, offering a foundation for further investigation into personalized obesity care.

In conclusion, this study exemplifies the dynamic interplay between genetic predispositions and gut microbiome with weight loss outcomes following sleeve gastrectomy. We observed significant associations between *Akkermansia* and EWL and evidence of association between multiple microbial pathways and polygenic scores that may inform personalized nutrition strategies. Future studies with extended follow-up will be essential to validate these results, establish causal relationships, and refine personalized treatment strategies to optimize long-term weight loss outcomes and metabolic health.

## Declaration of Generative AI and AI-assisted technologies in the writing process

During the preparation of this work, the authors used www.perplexity.ai, www.chatgpt.com, and https://consensus.app to improve the flow and readability of the text and to perform a complementary literature review to contextualize the findings of the study. After using these tools/services, the authors reviewed and edited the content as needed and take full responsibility for the content of the publication.

## Data Availability Statement

The microbiome sequence data used in this study were submitted to NCBI SRA under Bioproject accession number PRJNA1199984. The names of the repository/repositories and accession number(s) can be found in the article/Supplementary material. Supplementary materials provide individual-level data, processed gut microbiome abundances, and polygenic scores necessary to reproduce all results presented in the tables and figures of this study.

## Author Contribution

LT, RS, NS: Designed the study. GS, NS, SJ, DEA: Coordinated, monitored, and executed the study and data collection. IP, NS, SVK, KMM, SKS, SI: collected data, processed, curated, and cleaned the data and performed data analyses. IP, NS, SVK, SJ, DEA: wrote the manuscript. All authors reviewed and approved the manuscript.

