## Supplementary material for "Microbiome and Genetic Predictors of Weight Loss 12 Months Post Sleeve Gastrectomy: Insights from a Pilot Retrospective Cohort Study": Legends Supp Tables

### Supplementary Tables

**Table S1.** Table provides link to reference of database entry for the polygenic scores included in the analyses.

**Table S2.** The table provides metadata and demographics variables and health outcomes included in the analyses.

**Table S3.** Associations results obtained from regression models as described in the main text methods section. A positive Coefficient indicates a positive correlation between EWL and taxa abundance. n = sample size considered for the analysis.

**Table S4.** Associations results obtained from regression models as described in the main text methods section. A positive Coefficient indicates a positive correlation between EWL and pathway abundance. n = sample size considered for the analysis.

**Table S5.** The table provides abundance for genera reaching nominally significant association.

**Table S6.** The table provides abundance for microbial pathways reaching nominally significant association.

**Table S7.** Associations results obtained from regression models as described in the main text methods section. A positive Coefficient indicates a positive correlation between EWL and PGS. n = sample size considered for the analysis.

**Table S8.** The table provides PGS values for scores reaching nominally significant association.
