## Supplementary Figures for "Microbiome and Genetic Predictors of Weight Loss 12 Months Post Sleeve Gastrectomy: Insights from a Pilot Retrospective Cohort Study"

Inti Pedroso BSc PhD MBA [1 †], Shreyas V. Kumbhare MSc PhD [1 †], Shaneeta Johnson MD MBA FACS FASMBS ABOM [3], Karthik M. Muthukumar MS [1], Santosh K. Saravanan BTech [1], Carmel Irudayanathan BSc, MHRM[1], Garima Sharma MD MBBS[1], Lawrence Tabone MD, MBA, FACS, FASMBS [2], Ranjan Sinha MBA[1], Daniel E. Almonacid BSc PhD [1╓*] Nova Szoka MD, FACS, FASMBS [2╓*]

[1]Digbi Health, CA, USA

[2]Department of Surgery, West Virginia University, WV, USA

[3]Department of Surgery, Morehouse School of Medicine, GA, USA

### Supplementary Figures

**Figure S1.** provides a boxplots representing the distribution of EWL stratified by those that did (n=34) and did not (n=33) achieve successful weight loss after SG (EWL ≥ 50%).

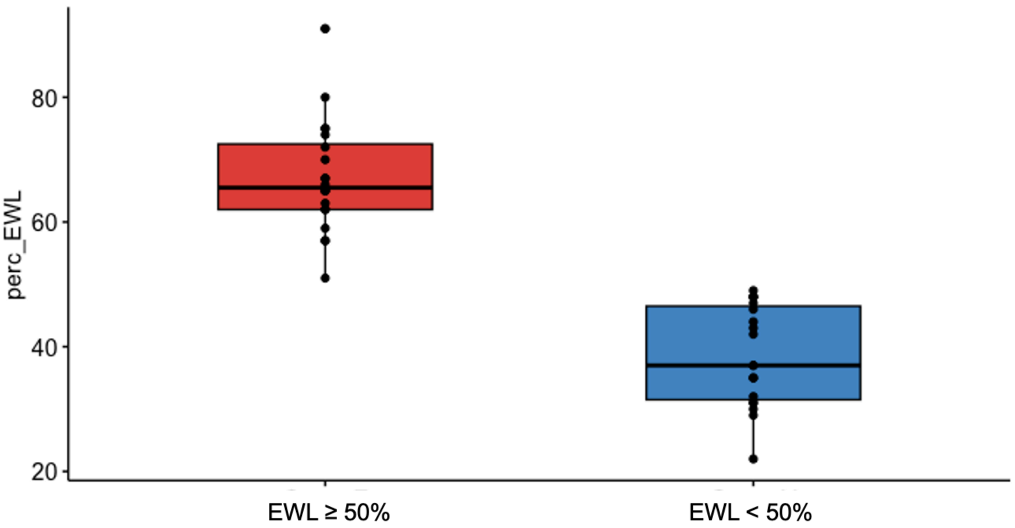

**Figure S2.** (A) Principal Component Analysis (PCA) of the fecal microbiota composition of patients with EWL ≥ 50% and EWL < 50%; p-value from the PERMANOVA based on Bray-Curtis dissimilarity. (B) Firmicutes/Bacteroidetes ratio between patients with EWL ≥ 50% (n=34) and EWL < 50% (n=33); p-value from Kruskal-Wallis sum rank test.

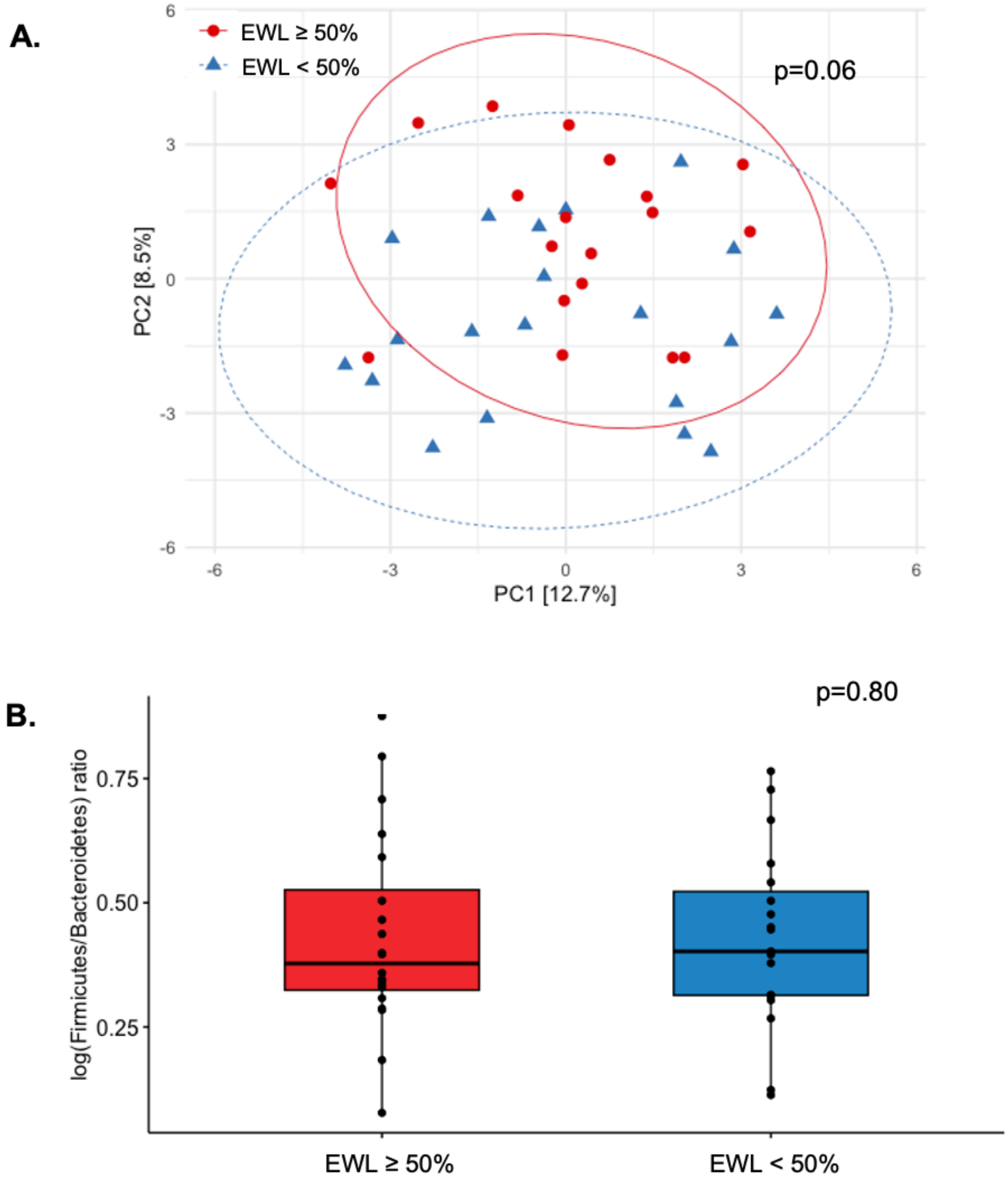

**Figure S3.** Box plot of abundance (y-axis) gut microbiome genus Akkermansia in individuals stratified by achieving or not achieving EWL ≥ 50% (x-axis). Violin and boxplot depict the sample abundance distribution and the points represent individual samples.

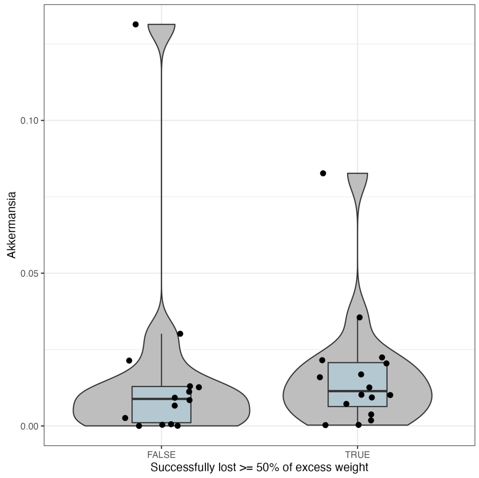


**Figure S4.** Box plot of abundance (y-axis) gut microbiome genus Family XIII UCG-001 in individuals stratified by achieving or not achieving EWL ≥ 50% (x-axis). Violin and boxplot depict the sample abundance distribution and the points represent individual samples.

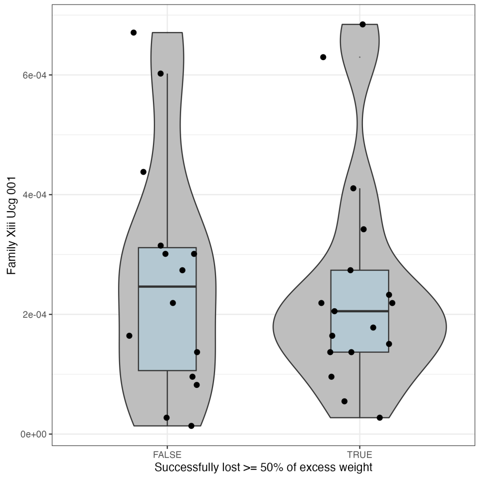

**Figure S5.** Box plot of abundance (y-axis) gut microbiome genus *Eubacterium hallii* group in individuals stratified by achieving or not achieving EWL ≥ 50% (x-axis). Violin and boxplot depict the sample abundance distribution and the points represent individual samples.

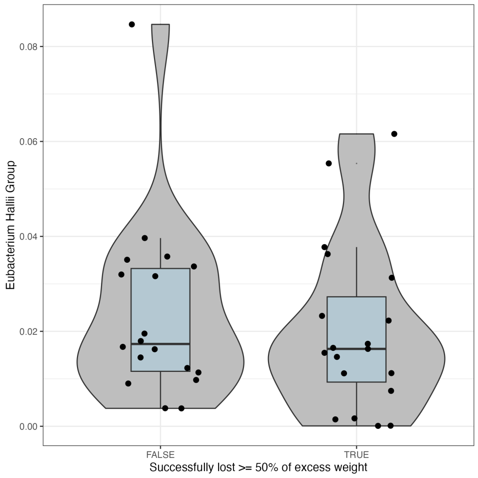


**Figure S6.** Box plot of abundance (y-axis) gut microbiome genus *Dorea* in individuals stratified by achieving or not achieving EWL ≥ 50% (x-axis). Violin and boxplot depict the sample abundance distribution and the points represent individual samples.

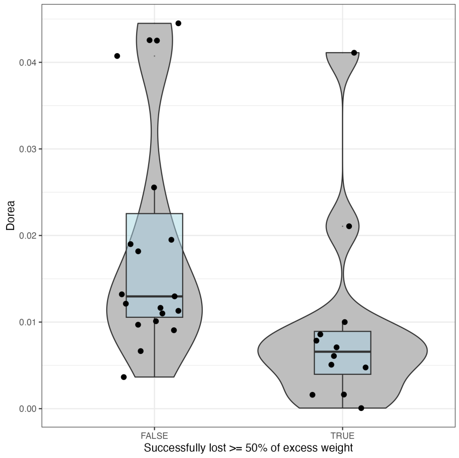

**Figure S7.** Box plot of abundance (y-axis) gut microbiome genus *Adlercreutzia* in individuals stratified by achieving or not achieving EWL ≥ 50% (x-axis). Violin and boxplot depict the sample abundance distribution and the points represent individual samples.

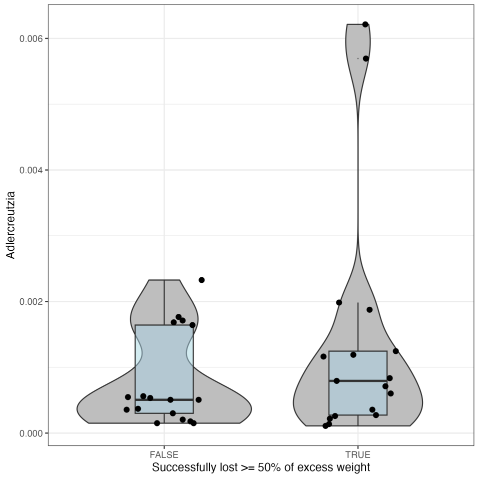


**Figure S8.** Box plot of abundance (y-axis) gut microbiome genus *Lachnospira* in individuals stratified by achieving or not achieving EWL ≥ 50% (x-axis). Violin and boxplot depict the sample abundance distribution and the points represent individual samples.

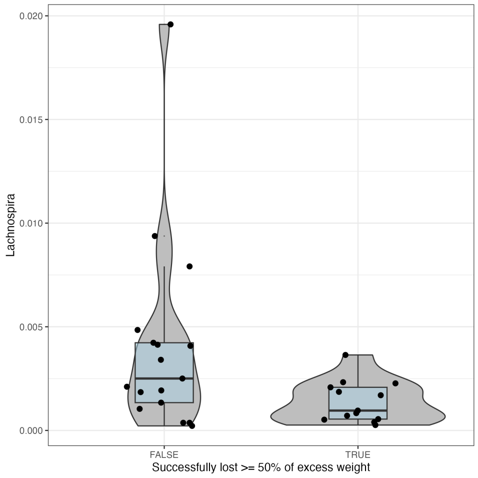

**Figure S9.** Box plot of abundance (y-axis) gut microbiome microbial pathway "MF0016: fucose degradation" in individuals stratified by achieving or not achieving EWL ≥ 50% (x-axis). Violin and boxplot depict the sample abundance distribution and the points represent individual samples.

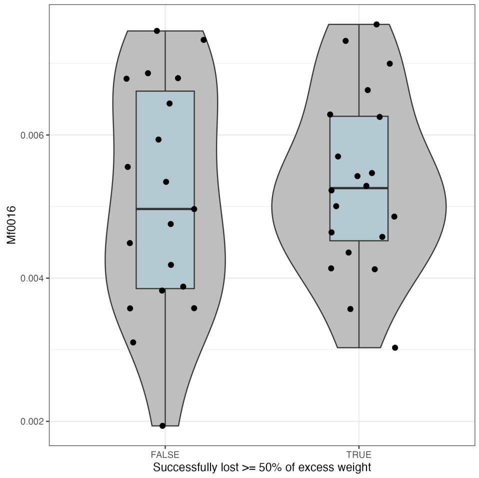


**Figure S10.** Box plot of abundance (y-axis) gut microbiome microbial pathway "MGB054: Propionate synthesis II" in individuals stratified by achieving or not achieving EWL ≥ 50% (x-axis). Violin and boxplot depict the sample abundance distribution and the points represent individual samples.

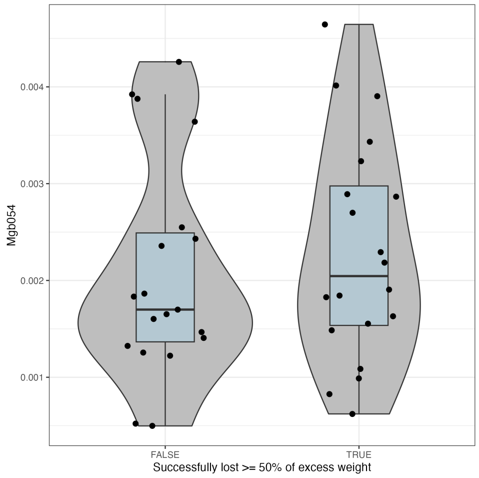

**Figure S11.** Box plot of abundance (y-axis) gut microbiome microbial pathway "MF0094: propionate production II" in individuals stratified by achieving or not achieving EWL ≥ 50% (x-axis). Violin and boxplot depict the sample abundance distribution and the points represent individual samples.

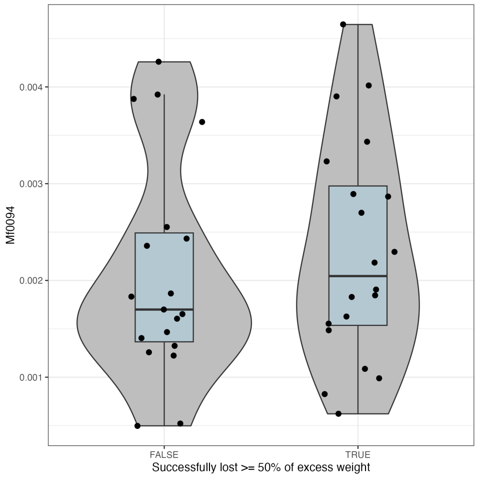


**Figure S12.** Box plot of abundance (y-axis) gut microbiome microbial pathway "MF0080: lactate consumption II" in individuals stratified by achieving or not achieving EWL ≥ 50% (x-axis). Violin and boxplot depict the sample abundance distribution and the points represent individual samples.

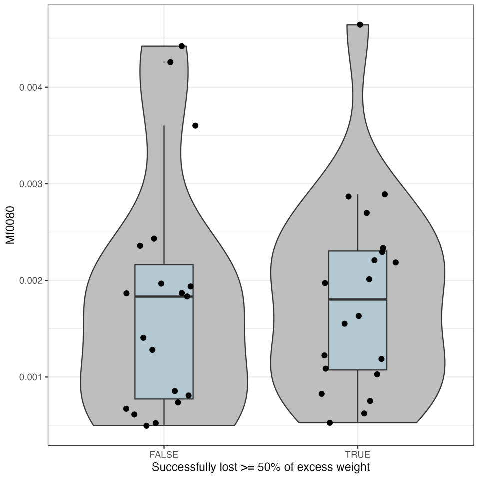

**Figure S13.** Box plot of abundance (y-axis) gut microbiome microbial pathway "MGB041: Menaquinone synthesis (vitamin K2) II (alternative pathway: futalosine pathway)" in individuals stratified by achieving or not achieving EWL ≥ 50% (x-axis). Violin and boxplot depict the sample abundance distribution and the points represent individual samples.

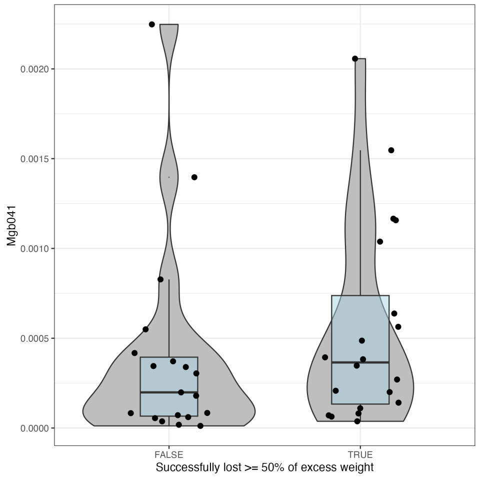


**Figure S14.** Box plot of abundance (y-axis) gut microbiome microbial pathway "MF0001: arabinoxylan degradation" in individuals stratified by achieving or not achieving EWL ≥ 50% (x-axis). Violin and boxplot depict the sample abundance distribution and the points represent individual samples.

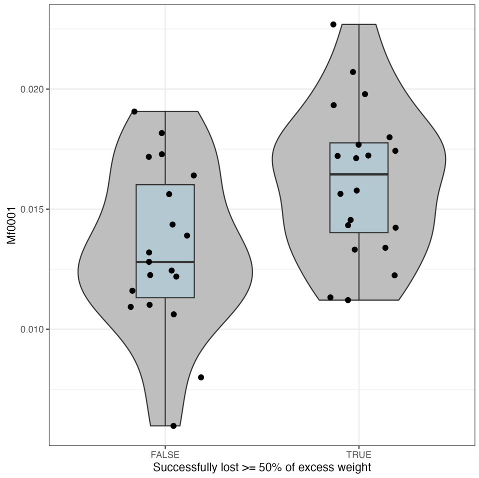

**Figure S15.** Box plot of abundance (y-axis) gut microbiome microbial pathway "MF0102: sulfate reduction (dissimilatory)" in individuals stratified by achieving or not achieving EWL ≥ 50% (x-axis). Violin and boxplot depict the sample abundance distribution and the points represent individual samples.

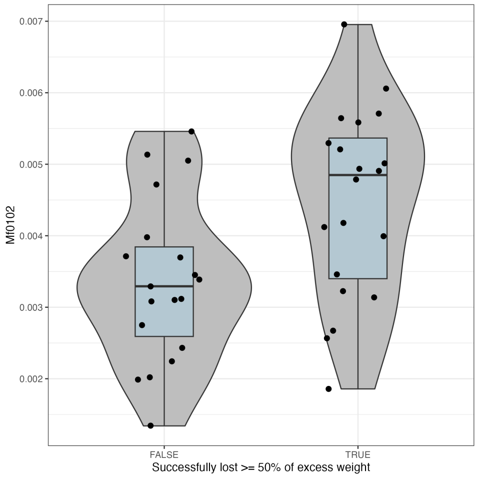


**Figure S16.** Box plot of abundance (y-axis) gut microbiome microbial pathway "MF0014: arabinose degradation" in individuals stratified by achieving or not achieving EWL ≥ 50% (x-axis). Violin and boxplot depict the sample abundance distribution and the points represent individual samples.

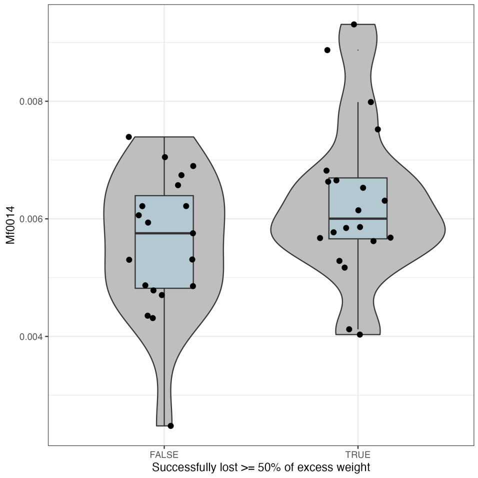

**Figure S17.** Box plot of abundance (y-axis) gut microbiome microbial pathway “MF0037: leucine degradation” in individuals stratified by achieving or not achieving EWL ≥ 50% (x-axis). Violin and boxplot depict the sample abundance distribution and the points represent individual samples.

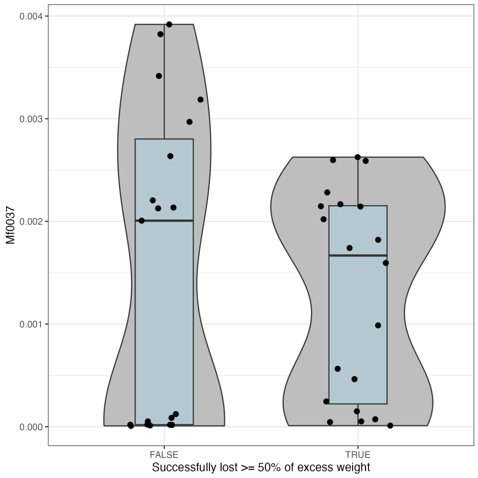


**Figure S18.** Box plot of PGS values (y-axis) of score “Vitamin D needs” in individuals stratified by achieving or not achieving EWL ≥ 50% (x-axis). Violin and boxplot depict the sample abundance distribution and the points represent individual samples.

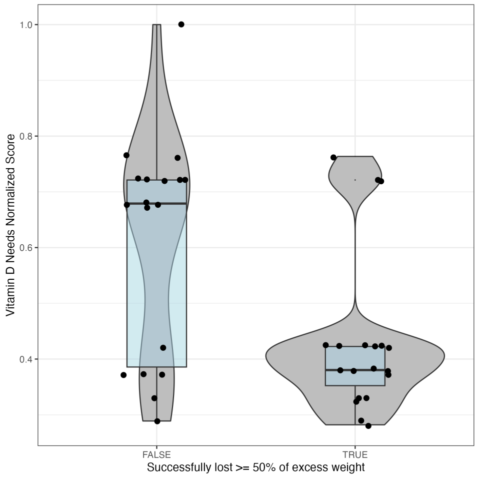

**Figure S19.** Box plot of PGS values (y-axis) of score “Vitamin K needs” in individuals stratified by achieving or not achieving EWL ≥ 50% (x-axis). Violin and boxplot depict the sample abundance distribution and the points represent individual samples.

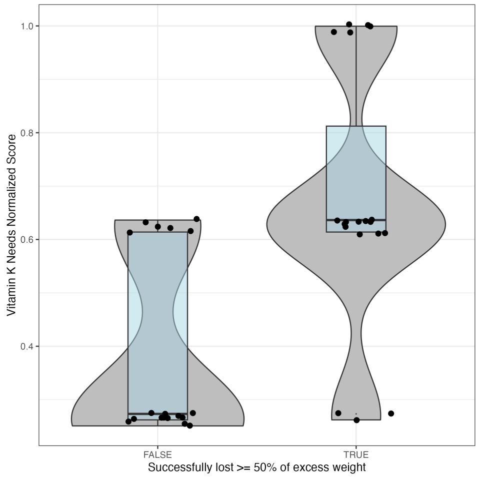


**Figure S20.** Box plot of PGS values (y-axis) of score “Major Depression” in individuals stratified by achieving or not achieving EWL ≥ 50% (x-axis). Violin and boxplot depict the sample abundance distribution and the points represent individual samples.

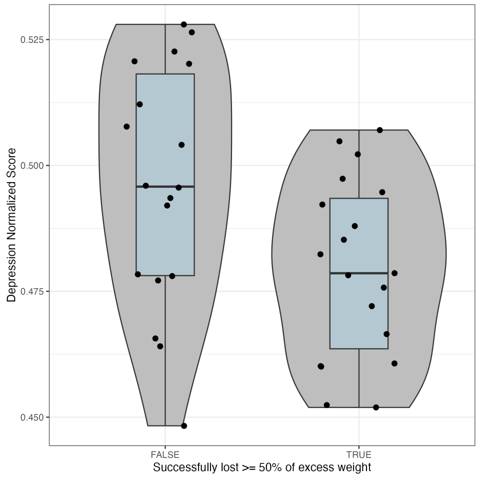

**Figure S21.** Box plot of PGS values (y-axis) of score "Psoriasis" in individuals stratified by achieving or not achieving EWL ≥ 50% (x-axis). Violin and boxplot depict the sample abundance distribution and the points represent individual samples.

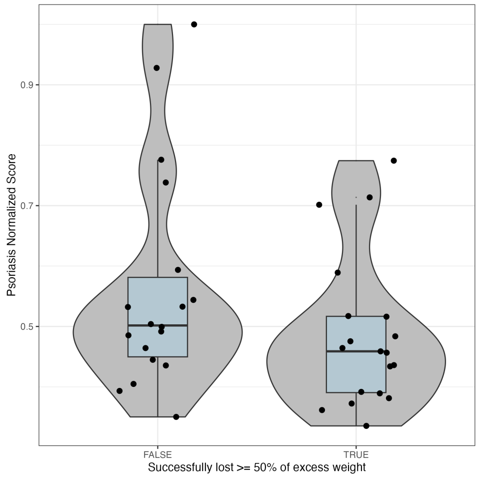


**Figure S22.** Box plot of PGS values (y-axis) of score "LDL Cholesterol" in individuals stratified by achieving or not achieving EWL ≥ 50% (x-axis). Violin and boxplot depict the sample abundance distribution and the points represent individual samples.

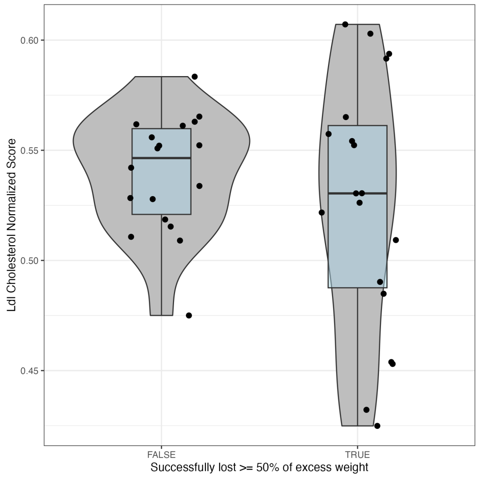

**Figure S23.** Box plot of PGS values (y-axis) of score "Protein Glycation" in individuals stratified by achieving or not achieving EWL ≥ 50% (x-axis). Violin and boxplot depict the sample abundance distribution and the points represent individual samples.

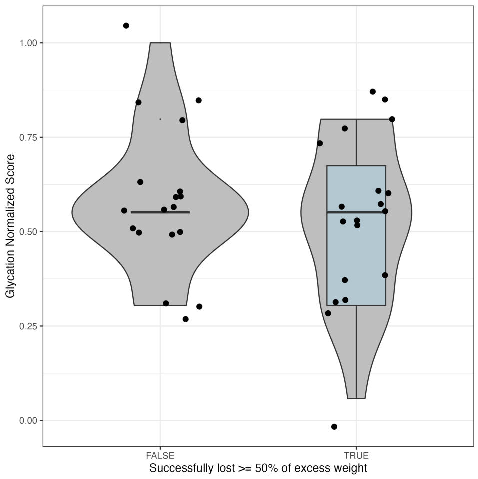
